# Nausea and Gastric Myoelectrical Activity are Influenced by Hormonal Contraception in Chronic Gastroduodenal Disorders

**DOI:** 10.1101/2024.04.20.24306132

**Authors:** Alexandria H Lim, Chris Varghese, Gabrielle Sebaratnam, Gabriel Schamberg, Stefan Calder, Armen Gharibans, Christopher N Andrews, Charlotte Daker, Daphne Foong, Vincent Ho, Michelle R Wise, Greg O’Grady

## Abstract

**Background:** Chronic gastroduodenal disorders such as chronic nausea and vomiting syndrome (CNVS), functional dyspepsia (FD) and gastroparesis, are more prevalent among young women, many of whom are hormonal contraception users. We aimed to evaluate the effects of hormonal contraception on symptom severity and gastric myoelectrical activity in people with chronic gastroduodenal disorders.

**Methods:** This analysis was conducted on a large international cohort of patients who met Rome IV criteria for CNVS or FD and had undergone body surface gastric mapping using Gastric Alimetry (Alimetry, New Zealand). Symptoms were continuously reported on 0-10 Likert scales using a validated symptom logging app.

**Results:** 127 people were included: 43 females using hormonal contraception, 30 premenopausal females not using hormonal contraception, 30 postmenopausal females, and 24 males. Premenopausal females who used hormonal contraception had higher nausea scores than non-users (3.80 [IQR 2.00-5.42] vs 2.25 [0.20-4.43]; p<0.05), particularly when using the combined oral contraceptive pill (COCP) with hormone-free intervals compared to continuous use (5.20 [4.30-6.00] vs. 2.40 [1.70-3.80], p=0.02). Premenopausal women were more symptomatic than postmenopausal women and men (p<0.001). The Principal Gastric Frequency was higher in hormonal contraception users (median 3.1 cpm [IQR 3.00-3.30] vs. 3.00 cpm [2.90-3.10], p<0.001), and highest in users of progestogen-only formulations (p<0.02).

**Conclusion:** Women with gastrointestinal disorders on hormonal contraception experience increased nausea in comparison to non-users of hormonal contraception, with substantial variation in nausea severity dependent on contraceptive type. Hormonal contraception users also demonstrated modified gastric electrophysiology. Women with chronic gastroduodenal symptoms should be asked about their use of hormonal contraception and non-hormonal contraceptive alternatives trialled as a means to reduce symptoms.

## Introduction

Chronic gastroduodenal disorders are poorly understood (1–3) and more commonly affect young female patients (4–8). The underlying cause of their disproportionate symptom burden is unknown. One contributing factor may be hormonal contraception, which has been clearly linked with an increased risk of gastrointestinal side effects in healthy women, including nausea and bloating (9,10). The relationship between female sex hormones and gastric symptoms may provide valuable insights towards understanding mechanisms underlying chronic gastroduodenal symptoms.

Hormonal contraception is a popular form of birth control, with over 150 million users worldwide receiving synthetic mimics of progesterone and estrogen (11–14). Hormonal interactions in the gastrointestinal tract have been proposed to contribute to gastric disorders (15,16). Symptoms have been seen to vary with hormonal fluctuations in the menstrual cycle in patients with chronic unexplained gastroduodenal symptoms (17–19). The gastric electrical system also appears sensitive to the administration of exogenous progesterone and estrogen, inducing dysrhythmic activity (20). Hormonal contraception could induce similar effects, but until recently it has been challenging to reliably and non-invasively assess gastric electrophysiology and gastric symptoms in relation to contraception use at scale.

Body surface gastric mapping (BSGM) is a new non-invasive diagnostic test for assessing gastric function (21–23). Validated metrics, determined by simultaneous gastric mapping and time-of-test symptom analyses, provide a standardised system to assess gastric electrophysiology (24–26). The aim of this study was therefore to evaluate the impact of hormonal contraception on nausea in patients with chronic gastroduodenal symptoms using a commercial BSGM system.

## Methods

A total of 129 patients were evaluated from a collaborative prospective database containing participants from three countries (Auckland, New Zealand; Western Sydney, Australia; Calgary, Canada). Ethical approval was granted for the protocol under the registrations AHREC 1330 (Auckland Health Research Ethical Committee, REB19-1925 (University of Calgary) and H13541 (Human Research Ethics Committee, Western Sydney). All patients provided written informed consent.

### Study Population

People aged ≥18 years were eligible for inclusion if they demonstrated a gastroduodenal symptom burden sufficient to meet the Rome IV Criteria for chronic nausea and vomiting syndrome (CNVS) or functional dyspepsia (FD), irrespective of gastric emptying status. Exclusion criteria were: incomplete test record, poor test quality (artifact >50% or poor impedance), current gastrointestinal (GI) infection, history of inflammatory bowel disease (IBD), GI malignancy, previous GI surgery, regular cannabis use, pregnancy, or metabolic or neurological disease known to affect gastric function, with the exception of diabetes.

Patients were categorised into four groups based on self-reported age, sex and contraception usage. Contraceptive status was self-reported by patients on the day of testing. Premenopausal women were considered hormonal contraception users if they were on any form of contraception that administered exogenous progesterone and/or estrogen, including oral contraceptive pills, depo provera, progestogen implants (depot medroxyprogesterone acetate; Jadelle) and levonorgestrel intrauterine system (IUS). Non-users of hormonal contraception included those using barrier methods and copper intrauterine devices (IUD), or no form of contraception. Postmenopausal women self-reported menopause hormone therapy (MHT) use. No transgender individuals were included in this study.

### Study Procedure

Participants fasted overnight before undergoing non-invasive BSGM using a high-resolution 64-electrode array connected to a wearable reader device (Gastric Alimetry^TM^, Alimetry, New Zealand) (22). Following a 30-minute fasting recording, participants consumed an energy bar (Clif Bar^™^ Nutrition Bar Chocolate Chip; 250 kcal, 5 g fat, 45 g carbohydrate, 10 g protein, 7 g fibre; Clif Bar & Company, Emeryville, CA) and nutrient drink (Ensure^™^, 230 kcal, 230 mL; Abbott Nutrition, Chicago, IL) and continued recording for a further 4 hours postprandially, per a standardized testing methodology (21,27). Continuous symptom monitoring was achieved using a validated symptom logging app, prompting subjects every 15 minutes to rate their nausea, bloating, upper gut pain, heartburn, stomach burn and excessive fullness on 0-10 visual analogue Likert scales (0 indicating no symptoms; 10 indicating the worst imaginable extent of symptoms) (24). Early satiation was recorded directly after meal consumption. All individual symptom scores were used to calculate the “Total Symptom Burden Score” over the duration of the test (24). Further detail on the standard Gastric Alimetry BSGM protocol is described extensively in other publications (22,28,29).

### Body Surface Gastric Mapping Metrics

The four spectral metrics reported by the Gastric Alimetry system were employed (25): Principal Gastric Frequency (PGF; the sustained frequency associated with the most stable gastric myoelectrical activity; normative interval 2.65 - 3.35 cpm), BMI-adjusted amplitude (normative interval 22 - 70μV), the Gastric Alimetry Rhythm Index (GA-RI; quantifying the extent to which activity is concentrated within a narrow frequency band over time relative to the residual spectrum; normative interval ≥0.25), and Fed:Fasted Amplitude Ratio (ff-AR, normative interval ≥1.08). A detailed description and validation for each metric and their reference intervals are detailed in the supplementary methods and elsewhere (25,26).

### Statistical Analysis

The primary outcome of this study was the relationship between hormone contraceptive use and nausea. Secondary outcomes were relationships between different modes of contraception with gastrointestinal symptoms and gastric myoelectrical activity. All analyses were performed using R (Version 4.2.3, R Foundation for Statistical Computing, Vienna, Austria). Non-parametric data are presented as the median and interquartile range (IQR). BSGM metric and symptom comparisons were made using Kruskall-Wallis and Wilcoxon signed-rank tests. Age and BMI adjustments were performed using generalised multivariable linear regression models. Post-hoc analyses were adjusted with Benjamini-Hochberg corrections.

## Results

Patients were from New Zealand (97/129, 76%), Australia (24/129, 18%) and Canada (8/129, 6%). Overall, 105/129 (81%) participants were female. Women on hormonal contraception were younger than women not on hormonal contraception (25 (20–29) vs. 31 (24–41), p = 0.007; see **Table 1**).

**Table 1:** Summary Baseline Characteristics.

|  |  | Users of<br>Hormonal<br>Contraception | Non-users of<br>Hormonal<br>Contraception | Post-Menopausal | Male |
| --- | --- | --- | --- | --- | --- |
| n (%) |  | 44 | 30 | 31 | 24 |
| Age (median) |  | 25 | 31 | 63 | 32 |
| BMI |  | 22.4 | 23.8 | 22.5 | 23.7 |
| Diagnosis<br>(n, (%)) | CNVS Only | 0 (0%) | 2 (6.7%) | 0 (%) | 0 (0%) |
|  | FD Only | 1 (2.3%) | 9 (30%) | 15 (48%) | 12 (50%) |
|  | Both | 43 (98%) | 19 (63%) | 16 (52%) | 12 (50%) |
| Contraceptive<br>Type | COCP with<br>HFI | 9 (20%) |  |  |  |
|  | Continuous<br>COCP | 7 (16%) |  |  |  |
|  | POP | 6 (14%) |  |  |  |
|  | Depo Provera<br>(DMPA) | 3 (6.8%) |  |  |  |
|  | Levonorgestrel<br>IUS | 17 (39%) |  |  |  |
|  | Other | 2 (4.5%) |  |  |  |

BMI = body mass index, CNVS = chronic nausea and vomiting syndrome, FD = functional dyspepsia, COCP = combined oral contraception pill, HFI = hormone-free interval, POP = progestogen only pill, IUS = intrauterine system

### Symptoms

Of the 129 participants, 90/129 (70%) met Rome IV criteria for both CNVS and FD, 37/129 (28%) for FD only and 2/129 (2%) for CNVS only. Excessive fullness was the most common symptom, experienced by 114/129 (88%), followed by bloating (102/129, 79%), upper gut pain (101/129, 78%), nausea (92/129, 71%), early satiation 91/129, 71%) and stomach burn (63/129, 49%). Belching was the most common symptom event registered during testing, experienced by 85/129 (66%) participants, followed by reflux (40/129, 31%) and vomiting (13/129, 10%).

Nausea scores were higher in hormonal contraception users compared to non-users of hormonal contraception (3.80 [IQR 2.00 to 5.42] vs. 2.25 [0.20 to 4.43], p = 0.04, p-adjusted = 0.05), postmenopausal women (0.05 [0.00 to 1.65], p <0.001, p-adjusted <0.001) and males (0.00 [0.00 to 1.80], p <0.001, p-adjusted <0.001) (**Figure 1A**). Post hoc corrections for multiple comparisons revealed nausea for hormonal contraception users remained higher than non-users of hormonal contraception (p-adjusted = 0.05), and both groups were higher than postmenopausal females and males (all p <0.02). There were no differences between hormonal contraception users and non-users for all other symptoms (all p >0.05) (**Figure 1C**).

**Figure 1:**
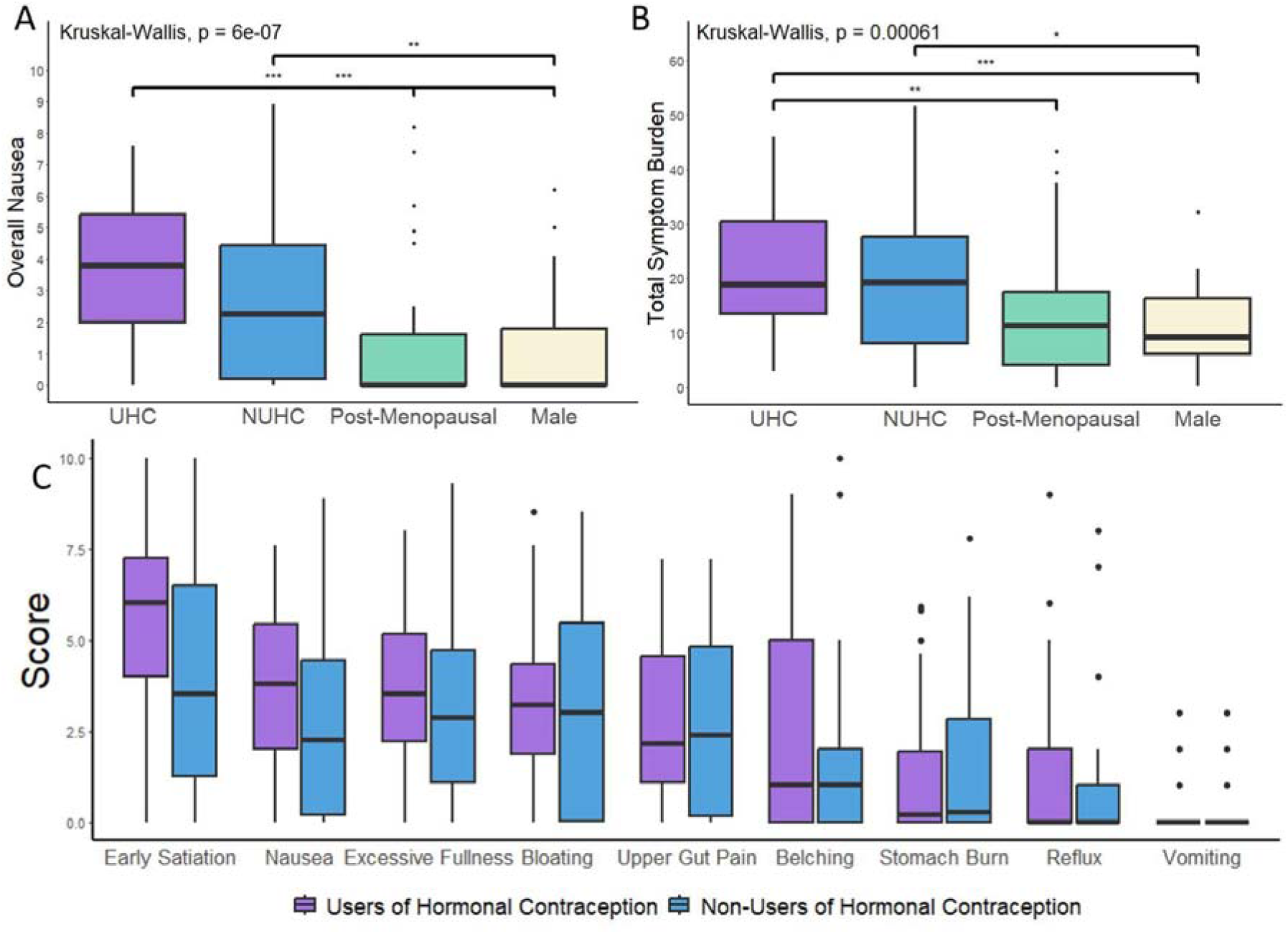
Symptom comparisons by use of hormonal contraception. A = Nausea, B = Total Symptom Burden, C = comparison of all symptoms between hormonal contraceptive users (UHC) and non-users (UNHC)

As demonstrated in **Figure 1B**, total symptom burden scores were higher in hormonal contraception users compared to males (median 18.80 [IQR 13.53 to 30.43] vs. 9.25 [IQR 6.18 to 16.28], p <0.001, p-adjusted <0.001). Non-users of hormonal contraception users also experienced higher total symptom scores than males (median 19.20 [IQR 8.15 to 27.68], p = 0.03, p-adjusted = 0.06), but total symptom burden score did not reach significance in premenopausal women by hormonal contraceptive use (p = 0.14, p-adjusted = 0.40).

#### Nausea by Contraceptive Type

Stratification by mode of contraception revealed significant differences in nausea scores in **Figure 2**. Premenopausal women who used the combined oral contraceptive pill (COCP) with hormone-free intervals (HFI) experienced more severe nausea than premenopausal women who used the COCP continuously (without HFI) (5.20 [4.30 to 6.00] vs. 2.40 [1.70 to 3.80], p = 0.023, p-adjusted = 0.06), premenopausal women who used the progestogen-only pill (POP) (3.25 [1.23 to 4.15], p = 0.026, p-adjusted = 0.07), and premenopausal women who did not use any form of hormonal contraception (2.25 [0.20 to 4.43], p = 0.02, p-adjusted = 0.06). Users of hormonal contraception had significantly higher nausea scores than postmenopausal women and men (all types p <0.01). Post hoc analyses by contraceptive type are provided in **Supplementary Material**.

**Figure 2:**
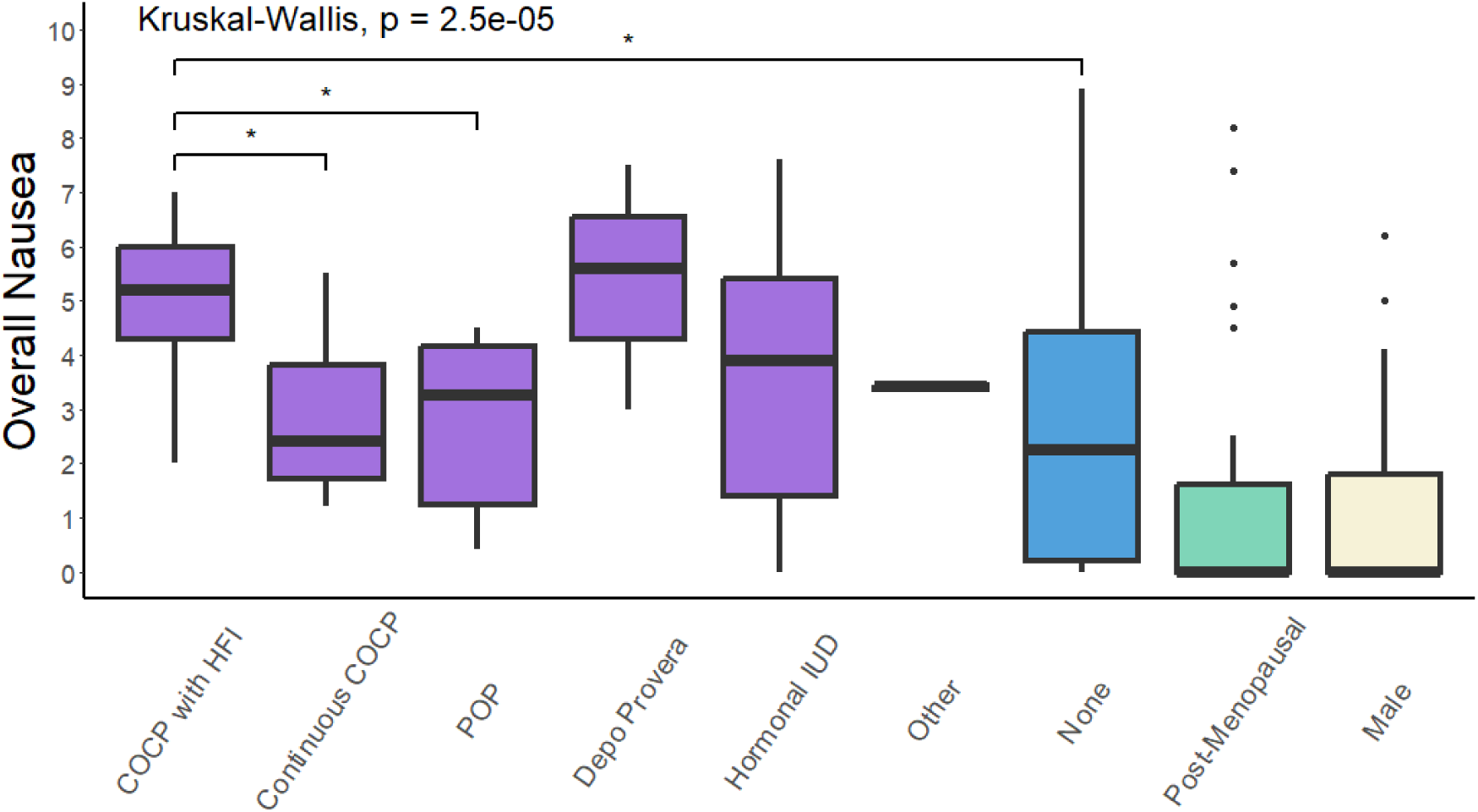
Comparing overall nausea scores with premenopausal women on hormonal contraception, stratified by type of contraception. COCP = combined oral contraceptive pill, HFI = hormone-free interval, POP = progestogen only pill, IUD = intrauterine device

#### Other Symptoms

Users of hormonal contraception demonstrated symptom differences when compared to postmenopausal women for overall bloating (3.20 [1.88 to 4.33] vs. 1.45 [0.00 to 3.58], p = 0.03, p-adjusted = 0.27), upper gut pain (2.15 [1.08 to 4.56] vs. 0.75 [0 to 2.23], p = 0.007, p-adjusted = 0.04), reflux (0.00 [0.00 to 2.00] vs. 0.00 [0.00 to 0.33], p = 0.02, p-adjusted = 0.14) and early satiation (6.00 [4.00 to 7.25] vs. 2.50 [0.00 to 6.00], p = 0.03, p-adjusted = 0.06).

Users of hormonal contraception also experienced higher symptom scores than males for upper gut pain (0.35 [0.00 to 2.08], p = 0.004, p-adjusted = 0.02), reflux (0.00 [0.00 to 0.00], p = 0.05, p-adjusted = 0.14), early satiation (2.00 [0.00 to 4.50], p = 0.002, p-adjusted = 0.01) and excessive fullness (3.50 [2.23 to 5.18] vs. 1.9 [0.85 to 2.70], p = 0.003, p-adjusted = 0.02).

Other symptoms analysed by mode of contraceptive are summarised in **Supplementary Material**.

#### Overall Trends

Users of hormonal contraception experienced the highest symptom burden across the majority of symptoms (belching, bloating, early satiation, excessive fullness, and nausea), although not all differences reached statistical significance. Non-users of hormonal contraception demonstrated lower symptom scores than hormonal contraception users but higher scores than postmenopausal women and men for bloating, early satiation and nausea (**Figure 3**).

**Figure 3:**
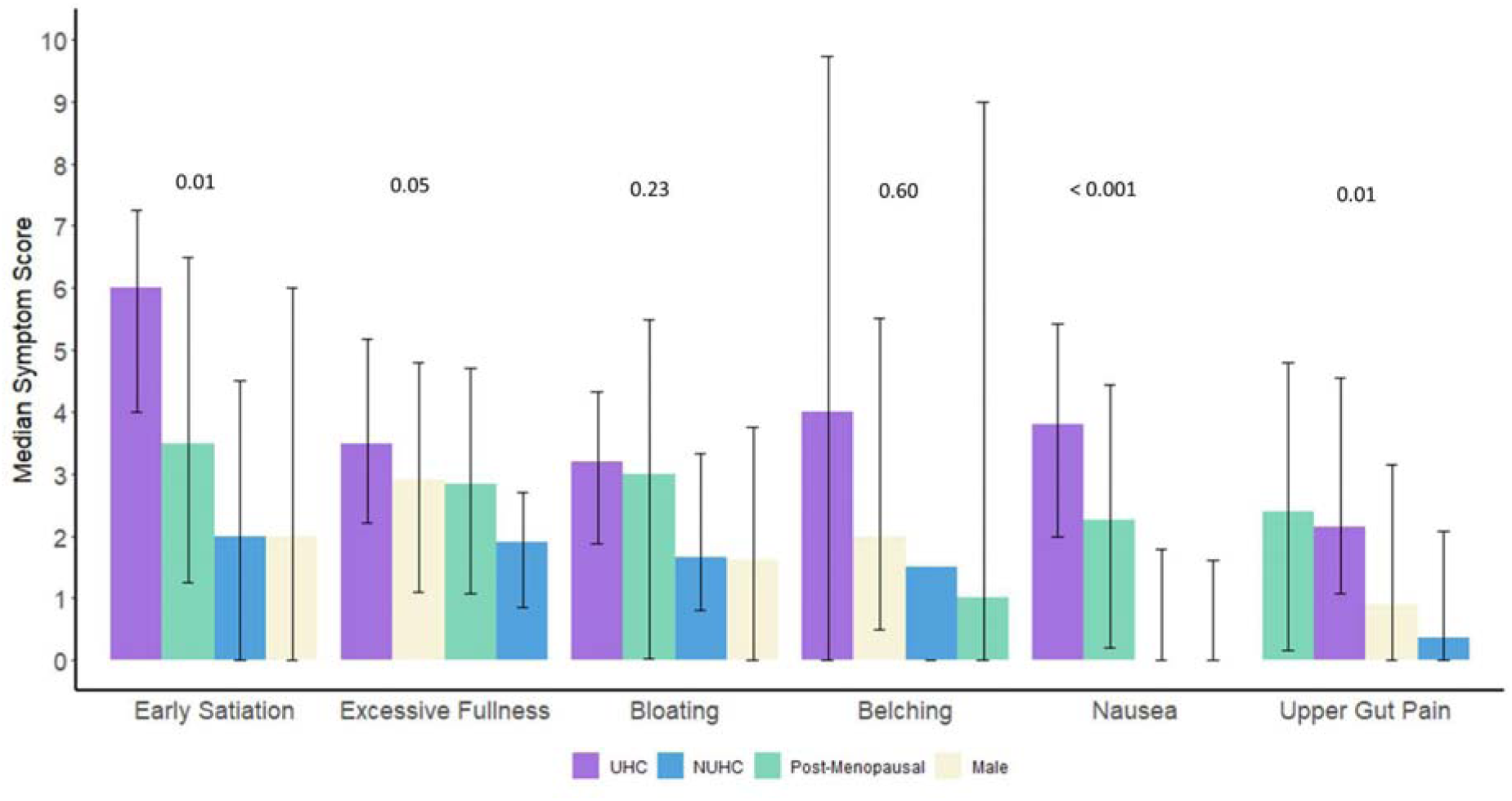
Median symptom scores by group across the 4.5-hour recording from highest to lowest score. The median score for vomiting and reflux was 0 for all groups, so were excluded from this figure. P values were calculated using the Kruskal-Wallis rank sum test.

### Gastric Mapping Metrics

Stratification by hormonal contraception use revealed several variations within BSGM metrics (**Figure 4A and 4B**). Users of hormonal contraception demonstrated higher PGF compared to non-users of hormonal contraception (median 3.10 cpm [IQR 3.00 to 3.30] vs. 3.00 cpm [2.90 to 3.10], p <0.001, p-adjusted = 0.004). Non-users of hormonal contraception users had higher BMI-adjusted amplitudes than males (31.90 [24.00 to 40.00] vs. 28.30 [19.57 to 33.23], p = 0.05, p-adjusted = 0.15) and hormonal contraception users had higher BMI-adjusted amplitudes (33.15 [27.93 to 40.90], p = 0.01, p-adjusted = 0.07) and GA-RI than males (0.42 [0.32 to 0.56] vs. 0.33 [0.26 to 0.43], p = 0.04, p-adjusted = 0.1). Postmenopausal females also had higher GA-RI than males (0.49 [0.32 to 0.58], p = 0.03, p-adjusted = 0.10). All groups demonstrated similar ff:AR (p = 0.39).

**Figure 4:**
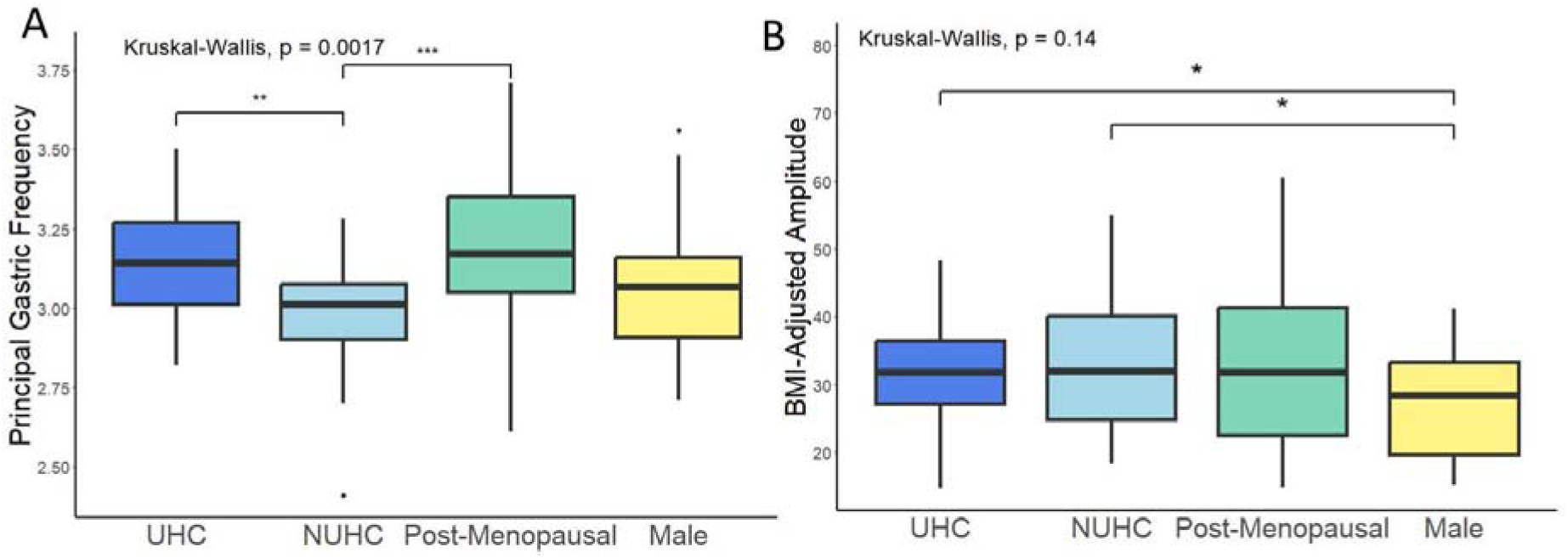
Comparative box plots for overall BSGM Metrics. A = Principal Gastric Frequency, B = BMI-Adjusted amplitude. Figures for the remaining metrics can be found in **Supplementary Figure 1**.

#### Principal Gastric Frequency by Mode of Contraception

When premenopausal females were categorised by contraception type, variations in PGF were also observed. As shown in **Figure 5**, females taking the POP had significantly higher PGF than females taking the COCP (regardless of hormone-free intervals) (with HFI; 3.28 [3.26 to 3.38] vs. 3.05 [3.03 to 3.07], p = 0.009), p adjusted = 0.1), (continuous; 3.14 [2.98 to 3.09], p = 0.003, p-adjusted = 0.1), females not on any form of contraception (3.01 [2.90 to 3.08], p <0.001, p-adjusted = 0.02), and males (3.07 [2.91 to 3.16], p = 0.02, p-adjusted = 0.2). Postmenopausal females also had significantly higher PGF than premenopausal females who did not use any form of hormonal contraception (3.17 [3.05 to 3.35] vs. 3.01 [2.90 to 3.08], p < 0.001, p-adjusted = 0.02).

**Figure 5:**
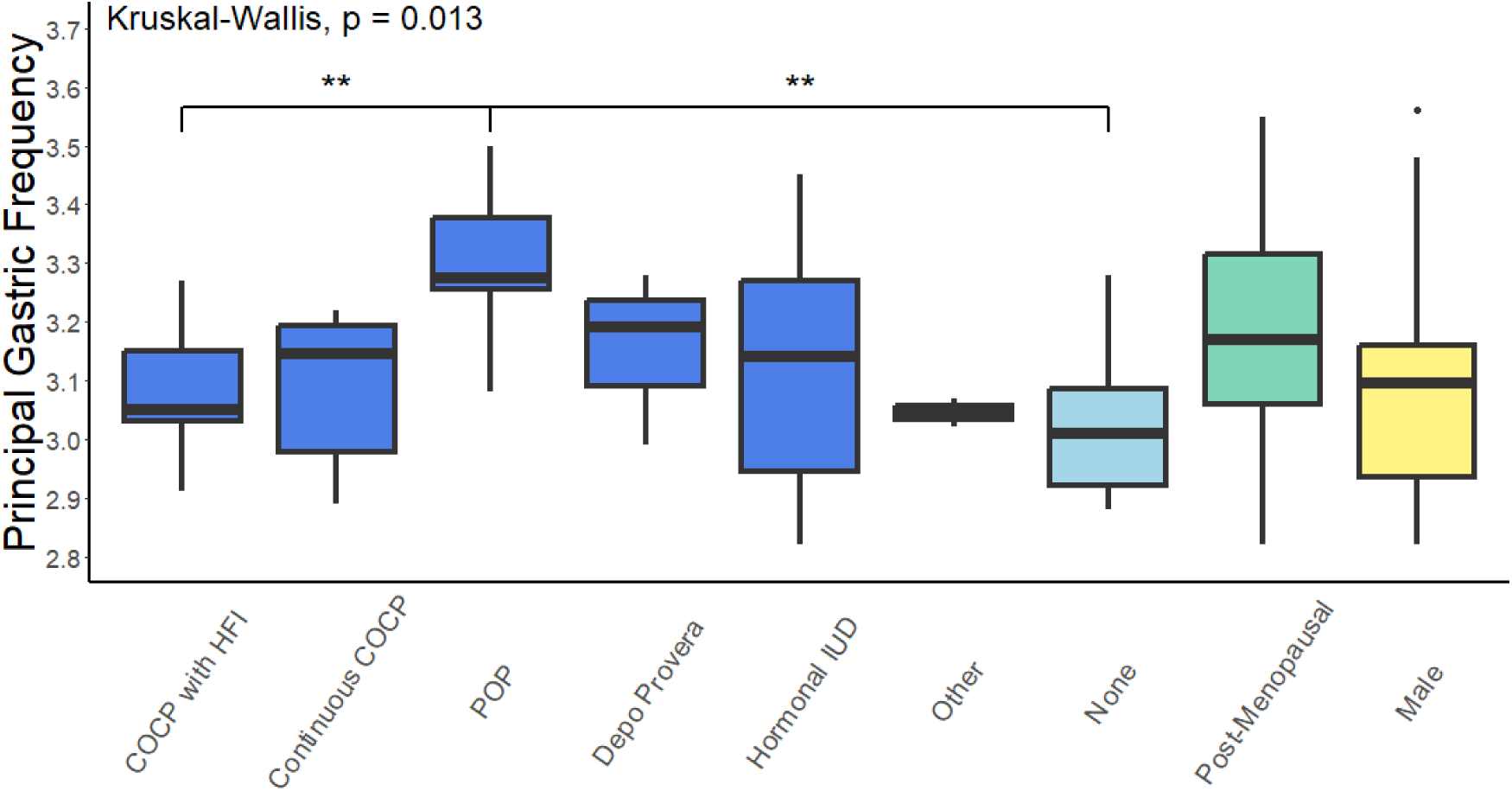
Principal Gastric Frequency by contraception type

#### Metrics Analysis - Age Adjustment

After adjusting for age, the predicted change in odds ratio for PGF remained significant between users of hormonal contraception and non-users of hormonal contraception (exp(β): 0.84; 95% CI 0.77, 0.92; p <0.001) and revealed an additional significant difference between hormonal contraception users and males (exp(β): 0.88; 95% CI 0.79, 0.99; p = 0.03). Comparisons between non-users of hormonal contraception and postmenopausal women fell below the threshold for significance (exp(β): 1.05; 95% CI 0.91, 1.21; p = 0.5).

## Discussion

This study demonstrates nausea is likely affected by hormonal contraception in chronic gastroduodenal disorders. The results confirm that premenopausal females experience a substantially higher burden of symptoms than postmenopausal and male patients. Importantly, we found that premenopausal women experience increased nausea in association with hormonal contraception use, with nausea severity being 1.7 times higher than in non-users. Substantial variation in nausea severity was also identified according to the specific mode of contraceptive used, with severity being 2.2 times higher for COCP users with hormone-free intervals compared to continuous COCP users. Hormonal contraception users and postmenopausal women also showed higher gastric frequencies compared to non-users, which also varied by mode of contraception, being highest in progesterone-specific formulations.

Gastroduodenal disorders are highly prevalent, with chronic nausea and vomiting disorders cumulatively affecting >1% of females, which is 2-3 times the rate reported in males (30,31). These demographic differences are specific to premenopausal women, with a recent population-based survey of >50,000 people from >25 countries showing that CNVS was identified in 1.3% of females aged 18-39, decreasing to 0.7% in ages 40-64, and 0.4% in ages 65+ (30). In a further study on patients with gastroparesis, Parkman et al. demonstrated that postmenopausal women generally experienced milder symptoms and relatively well-compensated disease compared to premenopausal women (OR 0.3) (4). Our results support a possible role for female sex hormones in explaining these discrepancies (18,32,33) and implicate hormonal contraception as a plausible contributing factor.

The role of hormonal contraception in gastroduodenal disorders appears to be relatively understudied. In gastroparesis, Verrengia et al. reported that non-oral contraceptive pill (OCP) users had cyclical variations in symptom severity that were lacking in OCP users, although overall symptoms appeared similar within their limited sample (n=20) (18). More recently, a large population-based (TriNetX) study comprising more than 3 million patients was reported in abstract form (34). The preliminary results indicated that within 5 years of starting COCPs, patients experienced a higher risk of nausea and vomiting, epigastric pain, and bloating; higher rates of investigations with endoscopy and gastric emptying studies; and higher rates of diagnoses with gastroparesis, functional dyspepsia, gastro-esophageal reflux disease, esophagitis and Barrett’s esophagus. While the full report of this study is still awaited, and confounding factors should be considered in a general population study, our data on nausea and hormonal contraception appears to show compatible findings.

While our study did not specifically address causality, a causal link can be reasonably postulated between hormonal contraception and nausea due to consistent and coherent evidence across adjacent literature, including temporal and dose-response relationships, as well as biological plausibility (35,36). For example, nausea and vomiting are commonly observed during fluctuations or surges in female sex hormones, as seen in menstrual cycling (18,37,38), pregnancy (39,40), as a side-effect of initiating hormonal contraception in healthy women (41,42); (43–46), and following the administration of emergency contraceptives (43–46). Symptoms have also been noted to fluctuate across the menstrual cycle in patients with gastroparesis, being heightened in the luteal phase (18). In addition, female sex hormones are well known to modulate several important aspects of GI function, including motility, intestinal permeability and mucosal immunity (32), with hormonal contraception also identified as a risk factor in other GI disorders, including inflammatory bowel disease (18,47,48). The incidence of gastric electrophysiological abnormalities has been shown to increase with higher doses of female sex steroids in animal studies (49,50). While the mechanisms of this dose-response relationship are not well established, central centres relevant to nausea expression may be involved, such as the chemoreceptive trigger zone (51).

In addition, our data demonstrated a surprising difference in nausea severity by the mode of hormonal contraception, further supporting a causal relationship (36), including the unexpected finding of a 2.2x higher nausea severity among COCP users with hormone-free intervals vs continuous COCP users. Hormone-free intervals result in greater fluctuations in serum estrogen and progesterone (52–54), and these fluctuations in circulating hormone levels could theoretically be responsible for exacerbating nausea in susceptible patients with pre-existing gastroduodenal disorders. This hypothesis is consistent with the fact that fluctuations in other nauseogenic stimuli can be more impactful on nausea and vomiting versus when stimulus levels are continuous or stable (55–57). Prospective cohort studies to evaluate nausea severity in women with gastroduodenal disorders using hormonal contraception are now desirable to confirm and extend these novel findings.

Based on the above rationale, the findings of this study are clinically significant. Chronic nausea disorders are challenging to treat (58,59). Women presenting with nausea should be asked about contraception use, and a trial of non-hormonal contraception can be considered to see if there is an improvement in symptoms. While healthcare professionals who prescribe contraception are aware that changes in contraception choice may become necessary if nausea occurs as a side effect, these changes are usually evaluated only around the time of prescription initiation; they may not currently be considered by gastroenterologists or for patients on long-term contraceptives who subsequently develop gastroduodenal disorders. The relatively simple intervention of changing contraceptive strategies could therefore be underutilised in GI practice and warrants further attention. In particular, prospective studies of patients with gastroduodenal disorders could be conducted to evaluate whether switching patients taking the COCP with hormone-free intervals to a continuous COCP regime or non-hormonal methods of contraception are effective strategies to reduce chronic symptoms and improve quality of life.

The changes in gastric electrophysiology observed in this study in association with hormonal contraception were important. The most notable change was in Principal Gastric Frequency, which was elevated with exogenous hormone delivery, particularly in formulations containing significant quantities of progestogen. This finding is consistent with a separate study recently performed by our group, which showed that similar elevations in gastric frequency occur normally during the luteal phase of the menstrual cycle in women not taking hormonal contraception when circulating progesterone is also elevated (38). Taken together, these data likely explain why females exhibit higher overall gastric frequencies than males in normative range datasets (25). While the changes in gastric frequency related to contraception use may appear modest, it should be noted that physiological homeostasis maintains PGF in a very tight range (2.65-3.35 cycles per minute) and thus a difference of 0.1 cpm, particularly after adjustment for age and multiple comparisons, reflects a relevant difference between the groups. It should be noted that the median PGF for all groups remained within a normal range (25), and the differences themselves are therefore unlikely to directly induce gastroduodenal symptoms; however, they may lead to small physiological increases in gastric mixing and emptying (38). In addition, while gastric dysrhythmias have previously been associated with both exogenous female sex hormone administration and nausea (20), dysrhythmias were not observed to be specifically associated with hormonal contraception use in our current study, as measured by the validated ‘Rhythm Index’ metric by the Gastric Alimetry System (see **Supplementary Figure 1**) (26). Regardless, the fact that contraceptive use has a measurable effect on gastrointestinal physiology and is associated with clinical symptoms is notable.

Some limitations of the current study are noted. The duration of contraceptive use was not available for this analysis, which may be relevant to the severity of nausea, given that patients may face greater side effects when first starting COCP (60). Participants using COCP may not have been administering uniform hormonal doses in this study as hormonal dosing between COCP formulations is variable. Subgroup analyses by hormonal type were restricted by the available sample size, and the novel nature of the subject limited the ability to inform a priori power calculations.

In summary, this study has revealed novel associations between hormonal contraception use, nausea severity, and gastric electrophysiology in patients with gastroduodenal disorders, which varied by the contraceptive type used. Contraception type is an important variable to consider in patients presenting with gastroduodenal disorders, and a trial of non-hormonal contraception should be considered as a potential management strategy.

## Data Availability

Data used for analysis will be made available upon reasonable request, conditional on ethical approvals.

## Acknowledgements

This research was conducted on behalf of the Body Surface Gastric Mapping (BSGM) consortium. We thank India Wallace and Gen Johnson for their invaluable research assistance.

## Supplementary Material

Post Hoc Analysis of Nausea by Contraceptive Type

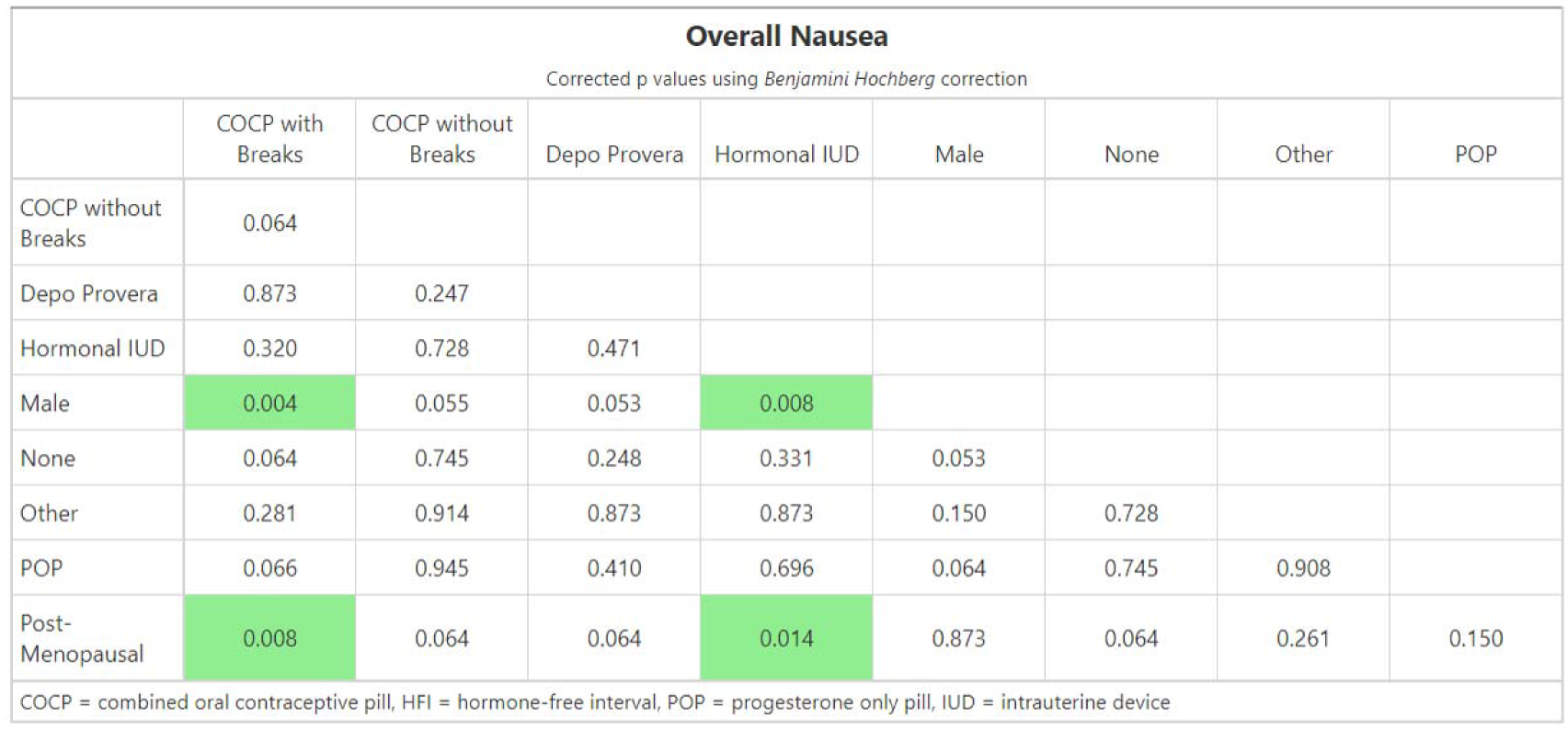

Analysis of Symptoms by Contraception Type

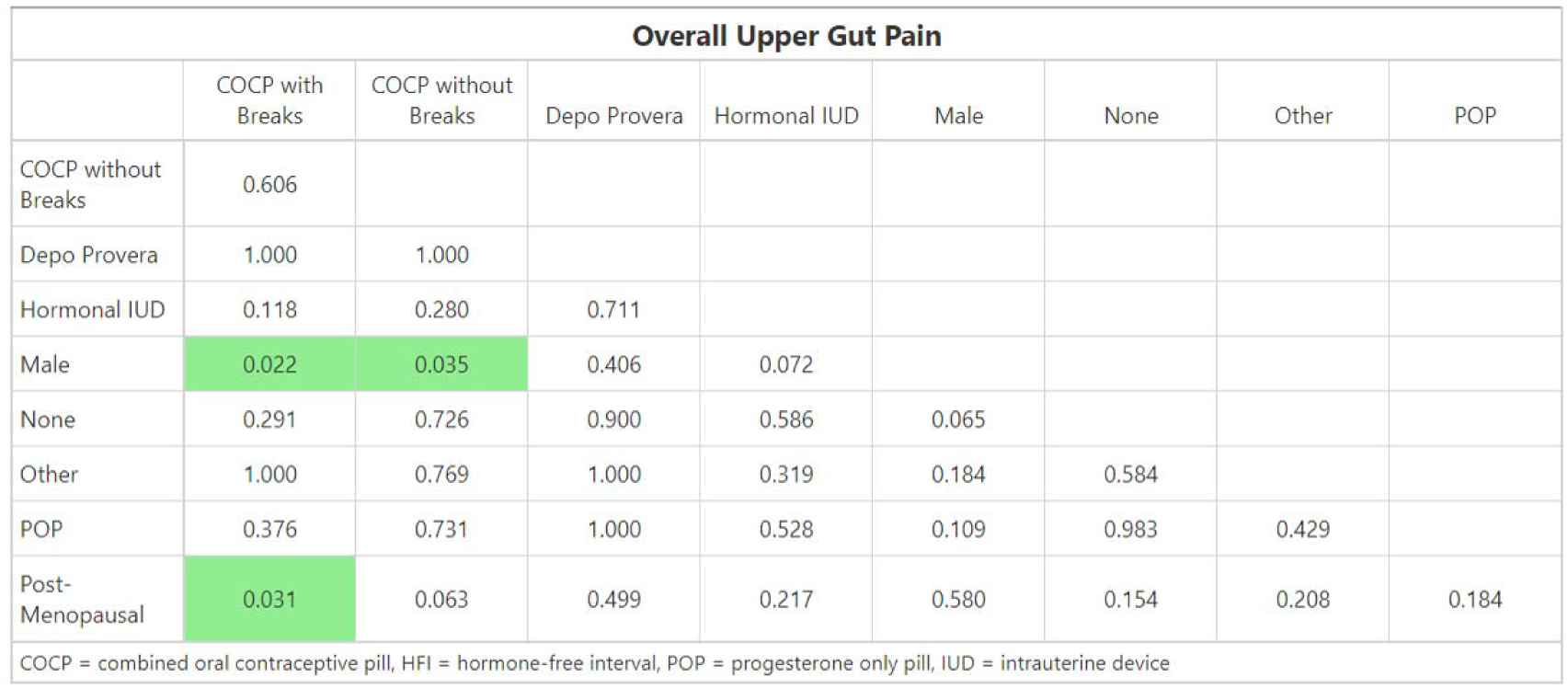
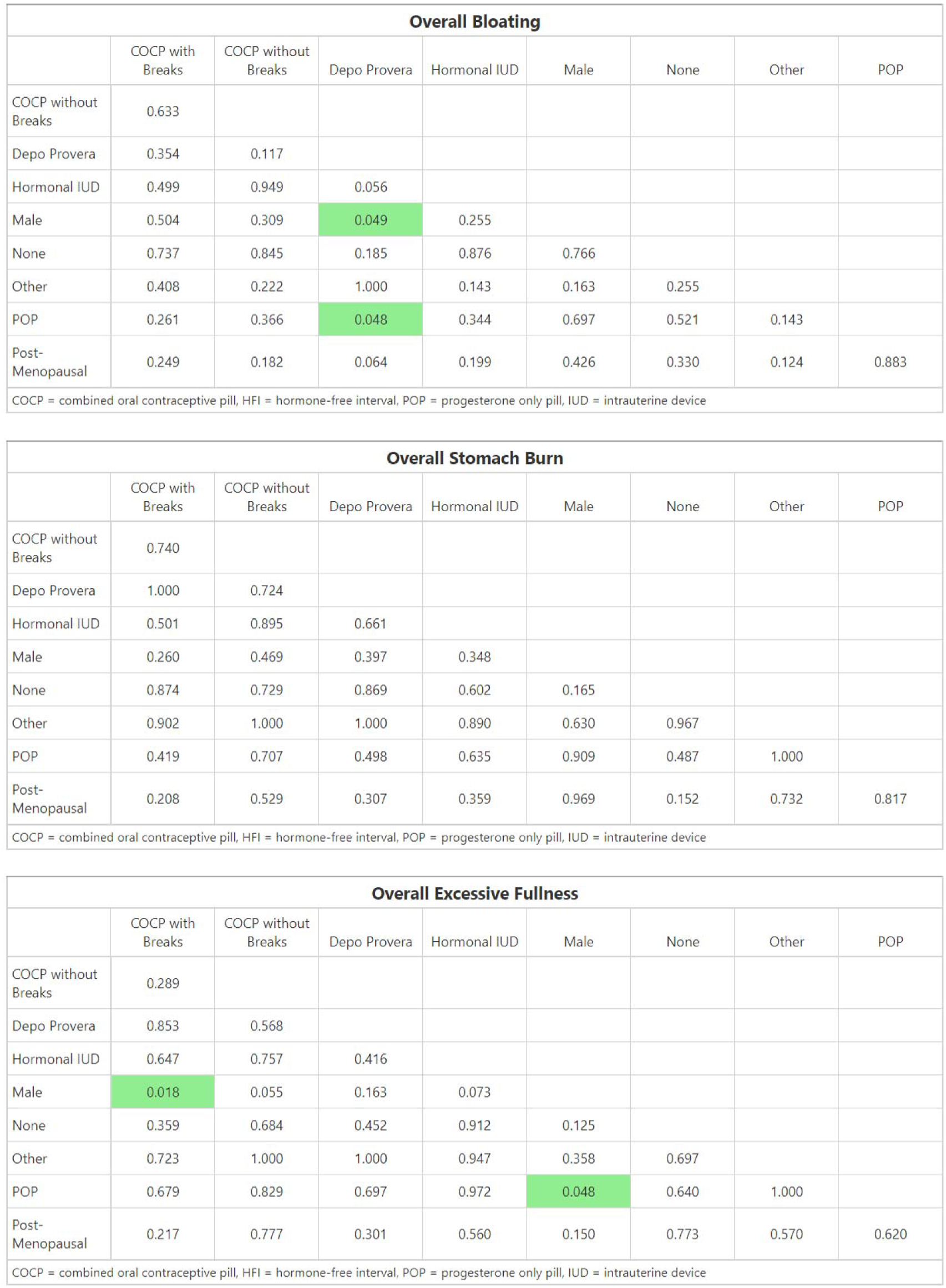
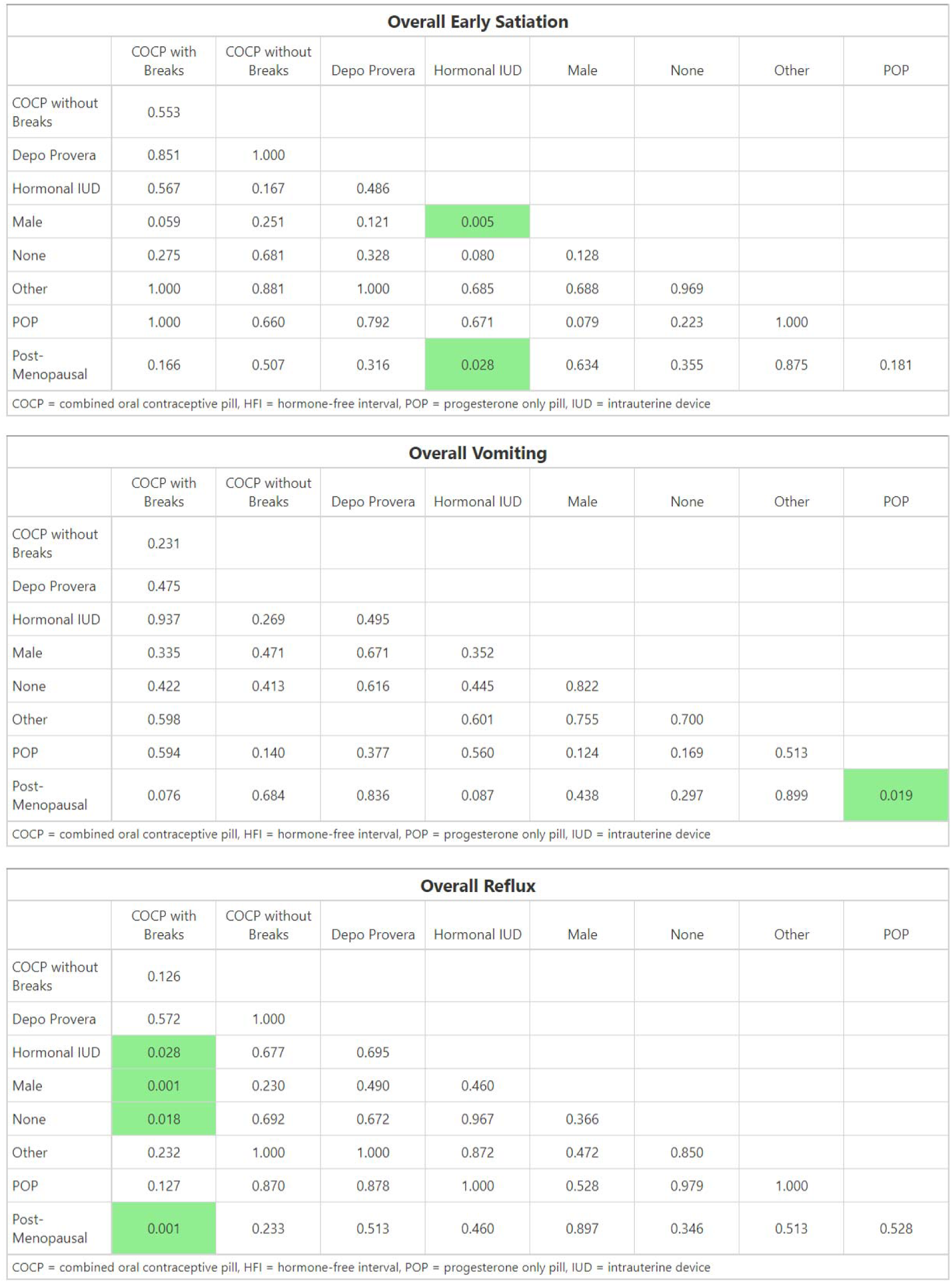
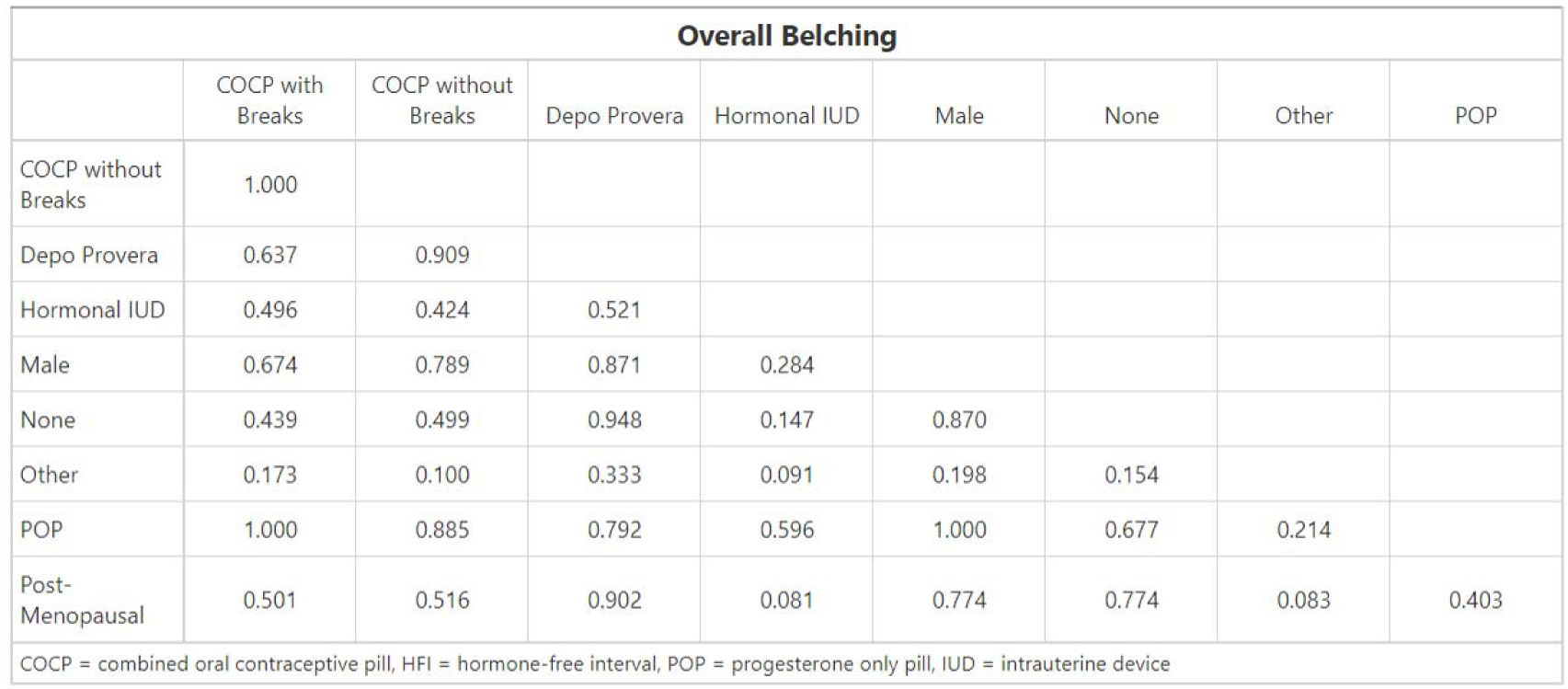

**Supplementary Figure 1:**
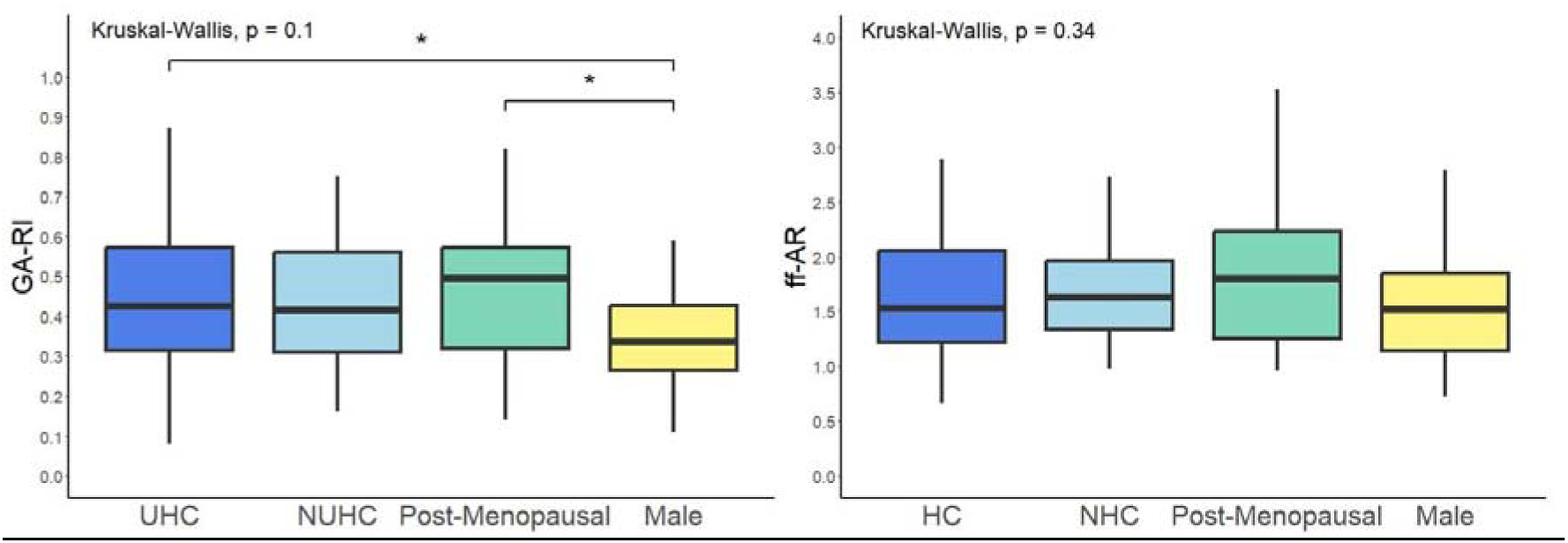
Boxplot comparison by contraception status for remaining BSGM Metrics. A = Gastric Alimetry Rhythm Index (GA-RI), B = Fed:Fasted amplitude ratio (ff-AR).

